# Diffusion MRI Is More Sensitive Than Tau PET and Regional Volume Loss in Corticobasal Syndrome

**DOI:** 10.64898/2026.07.24.26358494

**Authors:** Yuqi Tian, Ryota Satoh, Val J. Lowe, Keith A. Josephs, Jennifer L. Whitwell

## Abstract

**Background:** Corticobasal syndrome (CBS) is a neurodegenerative syndrome that often results from a 4-repeat tauopathy. Abnormalities on tau PET, structural MRI, and diffusion MRI (dMRI) have been observed in CBS, although no prior work has systematically compared the relative sensitivity of these modalities. In addition, the relationships between tau uptake, volume loss, and white matter degeneration remain incompletely understood.

**Objective:** To compare the sensitivity of tau PET, structural volume, and dMRI metrics to differentiate CBS from controls, and to characterize relationships between regional tau uptake, volume loss, and white matter tract abnormalities.

**Methods:** Thirty-two participants meeting criteria for possible or probable CBS and 35 healthy controls underwent standardized 3T dMRI, ^18^F-flortaucipir tau PET, and detailed neurologic assessment. Diffusion data were processed using complementary diffusion tensor, free-water, and Neurite Orientation Dispersion and Density Imaging (NODDI) pipelines, with tract-level metrics extracted using the Johns Hopkins University-Eve (JHU EVE) White Matter atlas. Regional gray matter volumes and flortaucipir standardized uptake value ratios (SUVRs) were calculated. Global tau PET uptake, represented by the first principal component (PC1), was removed in a secondary tau analysis. Modality-level discrimination was assessed using area under the curve (AUC) analyses. Cross-modality relationships were evaluated using regional correlations and covariate-adjusted models comparing volume- and tau-related predictors of white matter abnormalities.

**Results:** Relative to controls, CBS showed widespread higher mean diffusivity (MD) and lower intracellular volume fraction (ICVF), especially in sensorimotor and projection white matter pathways. Structural volume reductions were most prominent in precentral cortex and subcortical regions including putamen, thalamus, and pallidum, whereas tau PET abnormalities were weaker and less spatially extensive. MD showed the strongest overall discrimination between CBS and controls, followed by ICVF, structural volume, PC1-removed tau PET, and original tau PET. In targeted sensorimotor analyses, higher DTI-MD was most strongly associated with lower gray matter volume. PC1-removed sensorimotor tau uptake was also associated with higher sensorimotor DTI-MD, whereas original tau uptake and broader cortical or subcortical tau measures were not significant predictors.

**Conclusions:** White matter microstructural disruption is a dominant imaging signature of CBS and is more closely linked to gray matter volume loss than to measurable uptake on tau PET. dMRI, particularly MD and ICVF, may provide greatest sensitivity for detecting disease-related changes in CBS.

## Introduction

Corticobasal syndrome (CBS) is a progressive neurodegenerative disorder characterized by asymmetric parkinsonism, limb apraxia, cortical sensory loss, dystonia, myoclonus, language impairment, and executive dysfunction (Armstrong, Litvan et al. 2013). Clinically, CBS reflects asymmetric cortical and subcortical atrophy, with prominent degeneration of frontal and parietal regions and their associated motor networks (Boxer, Geschwind et al. 2006, Josephs, Whitwell et al. 2008). Although CBS is a clinical syndrome, rather than a pathologically defined entity, a substantial proportion of cases are associated with 4-repeat (4R) tauopathies, including corticobasal degeneration (CBD) (Jabbari, Holland et al. 2020).

Diffusion Magnetic Resonance Imaging (dMRI) offers a noninvasive means to probe white matter (WM) microstructural integrity across complementary tissue compartments (Tian and Whitwell 2026). Diffusion tensor imaging (DTI) measures such as mean diffusivity (MD) and fractional anisotropy (FA) capture broad microstructural disruption related to axonal loss, demyelination, or tissue disorganization (Basser and Pierpaoli 1996, Le Bihan, Mangin et al. 2001). More advanced modeling approaches, such as neurite orientation dispersion and density imaging (NODDI), provide estimates of intracellular volume fraction (ICVF), a putative marker of neurite density (Zhang, Schneider et al. 2012). Prior dMRI work in CBS has demonstrated DTI abnormalities in callosal, thalamic, and frontoparietal tracts, which correlate with key clinical features such as limb apraxia (Borroni, Garibotto et al. 2008, Tian, Ali et al. 2026).

Molecular biomarkers, particularly amyloid-β and tau, are central to biological characterization of CBS. Patients with evidence of both global amyloid-β and temporal lobe tau uptake on positron emission tomography (PET) fulfil criteria for biomarker-defined Alzheimer’s disease (AD) (Jack, Wiste et al. 2017, Jack, Therneau et al. 2019) and, indeed, tend to show AD characteristic patterns of hypometabolism on ¹⁸F-fluorodeoxyglucose (FDG) PET (Ghirelli, Goodrich et al. 2025). In contrast, studies of CBS without biomarker-AD tend to show patterns of tau PET uptake in the motor cortex (Ali, Whitwell et al. 2018), frontoparietal cortex, basal ganglia, and midbrain, with partial spatial overlap with cortical atrophy and FDG-PET hypometabolism (Nakano, Shimada et al. 2022). At the group level, tau PET uptake has been associated with dementia severity, supporting tau PET as a clinically relevant marker of disease burden, although not necessarily a complete explanation for clinical heterogeneity in CBS (Nakano, Shimada et al. 2022). More recent biomarker-based frameworks indicate that although most CBS patients are tau-positive, tau status alone shows limited ability to explain clinical heterogeneity or disease progression (Palleis, Bernhardt et al. 2026).

Despite growing use of dMRI and tau PET in CBS, two critical questions remain unresolved. First, it is unclear which imaging modality e.g. WM microstructure, regional brain volume, or tau PET provides the greatest sensitivity for detecting disease-related abnormalities. Second, the degree to which tau deposition and atrophy relate to WM injury, particularly within clinically relevant motor pathways, have not been directly compared within the same cohort. To address these gaps, we studied a cohort of 32 individuals with possible or probable CBS and 35 healthy controls who underwent tau PET, structural MRI-derived volume calculation, and multi-model dMRI. We excluded patients who had biomarker-confirmed AD to allow us to focus on CBS patients who are more likely to have an underlying 4R tauopathy. We aimed to (1) compare the discriminative sensitivity of tau PET, structural volume, and dMRI metrics, and (2) characterize cross-modality relationships between cortical tau uptake, regional volume loss, and WM microstructural abnormalities, with particular emphasis on motor-related pathways. To our knowledge, this is the first study to directly compare these imaging modalities and their interrelationships in CBS within a single, well-characterized cohort.

## Methods

### Participants

Participants were prospectively recruited by the Neurodegenerative Research Group (NRG) between January 2018 and December 2025 from the Department of Neurology, Mayo Clinic, Rochester, MN. We included all individuals who met research criteria for possible or probable CBS (Armstrong, Litvan et al. 2013). Because amyloid positivity alone may occur in older individuals and does not necessarily indicate that AD is the primary driver of the CBS phenotype, participants were excluded for biomarker evidence of AD only when both a global amyloid-beta PET meta-ROI and a temporal lobe tau PET meta-ROI were positive (Jack, Wiste et al. 2017, Jack, Therneau et al. 2019). Thus, this approach modified the Armstrong criteria by not excluding amyloid positivity in isolation. Thirty-two CBS participants met our inclusion criteria. These participants were matched by age and gender to 35 cognitively and motorically normal participants who did not have any complaints of cognitive, motor, or behavioral abnormalities and had a score of ≥23 on the Montreal Cognitive Assessment battery (MoCA) (Nasreddine, Phillips et al. 2005) and a score of 0 on the Hoehn and Yahr scale (Hoehn and Yahr 1967). All participants underwent the same standardized research MRI protocol and structured neurologic assessment. Twenty-nine out of 32 CBS participants, and 27 out of 35 controls, underwent tau PET scans.

The neurological assessment of the CBS participants included the Montreal Cognitive Assessment (MoCA) (Nasreddine, Phillips et al. 2005), the Movement Disorder Society sponsored revision of the Unified Parkinson’s Disease Rating Scale Part III (MDS-UPDRS III) (Goetz, Tilley et al. 2008), Progressive Supranuclear Palsy (PSP) Rating Scale (Golbe and Ohman-Strickland 2007), Frontal Assessment Battery (FAB) (Dubois, Slachevsky et al. 2000), Western Aphasia Battery Praxis subtest (WAB-Praxis) (Shewan and Kertesz 1980), PSP Saccadic Impairment Scale (PSIS) (Whitwell, Master et al. 2011), and the Test of Upper Limb Apraxia (TULIA) (Vanbellingen, Kersten et al. 2010), including left- and right-hand subscores when applicable. Disease-specific onset variables such as age at onset and duration from symptom onset to examination were also collected.

All participants provided written informed consent under protocols approved by the Mayo Clinic Institutional Review Board.

### MRI acquisition and preprocessing

All participants underwent a standardized MRI protocol on a 3 Tesla scanner (Magnetom Prisma; Siemens Healthineers, Erlangen, Germany). T1-weighted structural images were acquired using a 3D magnetization-prepared rapid gradient-echo (MPRAGE) sequence with the following parameters: repetition time (TR) = 2300 ms; echo time (TE) = 3.1 ms; inversion time (TI) = 945 ms; isotropic voxel size = 0.8 × 0.8 × 0.8 mm³; 240 sagittal slices. An atlas derived from the Mayo Clinic Adult Lifespan Template (MCALT) template was registered to each participant’s native T1 image space to enable ROI-based analyses. Atlas registration was performed using nonlinear transformation after an initial affine alignment, ensuring accurate correspondence between atlas regions and individual neuroanatomy. The resulting transformations were applied to propagate atlas labels into subject space. All registrations were visually reviewed to confirm appropriate alignment of cortical and subcortical structures. The T1-weighted images and derived tissue masks served as the anatomical reference for subsequent PET image co-registration and regional quantification. Regional gray matter (GM) volume estimates were obtained using MCALT atlas ROIs. Pons and dorsal mesopontine labels were excluded, resulting in 120 volume ROIs analyzed separately by hemisphere and adjusted for total intracranial volume (**Table S1**). Regional WM volume estimates were obtained using JHU EVE atlas ROIs (**Table S2**). To account for interindividual differences in head size, regional volumes were normalized by total intracranial volume (TIV) before statistical analysis. A sensorimotor GM volume composite was calculated from MPRAGE-derived volumes in predefined MCALT ROIs: bilateral precentral, postcentral, Rolandic operculum, supplementary motor area, and paracentral lobule (**Table S3**). Regional volumes were divided by TIV, standardized using the control-group mean and standard deviation, and averaged across sensorimotor ROIs. Lower composite z-scores indicate lower sensorimotor GM volume relative to controls. For predictor models in which higher values were intended to represent greater volume loss, the sensorimotor GM volume z-score was sign-inverted and labeled as sensorimotor GM volume loss.

Diffusion weighted imaging (DWI) sequence was acquired on the same 3T Siemens Prisma scanner with a 64-channel head-neck coil using a multi-shell high angular resolution diffusion imaging protocol (b = 0, 500, 1000, and 2000 s/mm^2; TE = 71 ms; TR = 3.4 s; 2 mm isotropic voxels; multiband factor = 3). Preprocessing followed a standard FSL and MRtrix3 workflow, including denoising, Gibbs ringing removal, motion and eddy-current correction with outlier replacement, bias-field correction, and quality control of geometric alignment and diffusion gradient tables. dMRI metrics were derived using multiple complementary modeling approaches. Fractional anisotropy (FA) and mean diffusivity (MD) were estimated from diffusion tensor imaging (DTI) using weighted least-squares fitting in DIPY (Garyfallidis, Brett et al. 2014). Free-water-corrected (FWE) FA and MD were generated using a two-compartment model in DIPY separating tissue and extracellular free-water signal. Neurite orientation dispersion and density imaging (NODDI) metrics, including intracellular volume fraction (ICVF), orientation dispersion index (ODI), and isotropic volume fraction (ISOVF), were estimated using AMICO (Daducci, Canales-Rodriguez et al. 2015). For NODDI-derived measures, tissue-weighted regional means were calculated to reduce bias from cerebrospinal fluid partial-volume effects. White matter diffusion metrics were extracted using the JHU EVE atlas (Table S2) after registration to each participant’s FA image; regions with less than 50% valid voxel coverage were excluded from analysis. Sensorimotor WM ROIs included bilateral corticospinal tract, precentral white matter, and postcentral white matter regions from the JHU-EVE atlas (Table S3).

### Tau PET

Tau PET imaging was performed using 18F-flortaucipir (AV-1451). Participants received an intravenous injection of approximately 370 MBq of 18F-flortaucipir, followed by a 20-minute PET acquisition beginning 80 minutes after injection. All scans were acquired on PET/CT scanners (GE Discovery 690XT, GE Discovery MI, or Siemens Biograph64 Vision 600). PET images were reconstructed using ordered-subsets expectation maximization with standard corrections and a 5-mm Gaussian post-reconstruction smoothing filter. The four 5-minute frames were motion-corrected and averaged to create a static image, which was rigidly registered to each participant’s T1-weighted MRI. Registration accuracy was confirmed by visual inspection (Satoh, Ali et al. 2024). Standardized uptake value ratio (SUVR) images were generated using median uptake in the cerebellar crus GM as the reference region. MCALT atlas ROIs were nonlinearly transformed into native subject space using MRI-based registrations and applied to the PET images. Mean 18F-flortaucipir SUVR was extracted from ROIs using combined gray- and WM masks.

To reduce the influence of a dominant global/shared tau signal, we generated an additional tau dataset with the first principal component (PC1) removed (Satoh, Ali et al. 2026). Principal component analysis was applied to the regional tau matrix across participants, and the variance explained by PC1 was regressed out to produce residual regional tau values for each subject. These PC1-removed tau measures were then used in parallel analyses to assess whether regional multimodal relationships were preserved after removal of the dominant shared tau pattern. Regional tau measures were analyzed as original SUVR z-scores and as PC1-removed SUVR z-scores.

Raw tau PET values extracted from the full MCALT atlas (Table S1) were used for the ROI ranking and display analyses shown in Figure 2 and Figure S1. Regional tau PET analyses were performed using selected cortical and subcortical MCALT atlas ROIs. The sensorimotor tau composite included precentral, postcentral, supplementary motor area, paracentral lobule, and Rolandic operculum ROIs. The subcortical tau composite included caudate, putamen, pallidum, and thalamus ROIs.

### Volume, dMRI and Tau

**Figure 1** illustrates the multimodal imaging data used in this study, with representative axial slices at the level of the lateral ventricles. Structural MRI includes T1-weighted MPRAGE images, from which GM and WM ROIs were derived. DWI with representative DTI and NODDI metrics were displayed. In addition, tau PET image demonstrating regional tau tracer uptake was displayed.

**Figure 1.**
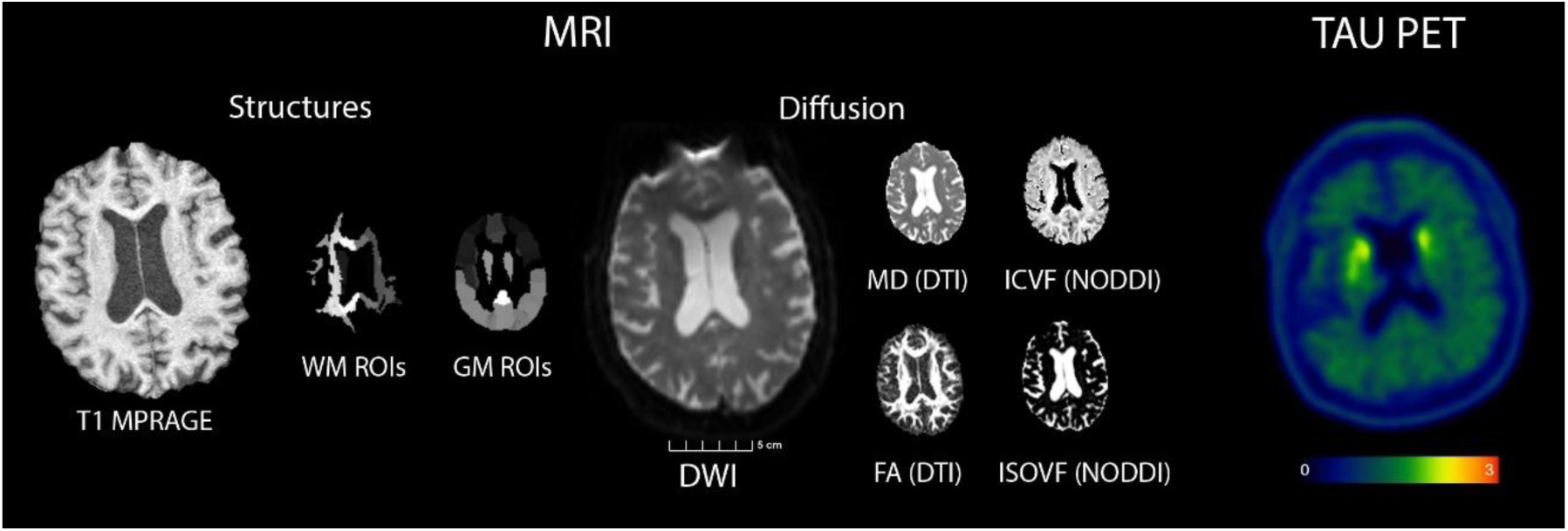
Representative axial images at the level of the lateral ventricles illustrate the multimodal imaging data used in this study. Shown are T1-weighted MPRAGE for GM and WM extraction, DWI (B0 shown in the figure) and representative DTI and NODDI metrics, and tau PET image demonstrating regional tau tracer uptake.

### Statistical analysis

All analyses were performed in Python using numpy, pandas, scipy, statsmodels, scikit-learn, and matplotlib. We first summarized cohort-level differences between participants with CBS and controls across regional imaging measures, including TIV-adjusted structural volumes, dMRI metrics, original tau PET SUVR z-scores, and PC1-removed tau PET SUVR z-scores. For these ROI-wise modality-level comparisons, regional effects were summarized using CBS-minus-control mean differences, Cohen’s d, Welch two-sample t-tests, and oriented univariate AUC. Benjamini-Hochberg false-discovery rate (FDR) correction was applied within each modality or metric family for ROI-wise group comparisons, and regions were ranked to identify the strongest CBS-control separation within each metric. Differences in AUC between modality-level sensorimotor summary measures were assessed using stratified nonparametric bootstrap resampling of subjects within diagnostic group with 10,000 iterations. Two-sided bootstrap p values were calculated from the distribution of paired AUC differences and adjusted using the Benjamini-Hochberg procedure.

We then examined targeted participant-level associations among sensorimotor volume, dMRI, original tau PET, and PC1-removed tau PET z-scores using two-sided Pearson correlations and covariate-adjusted ordinary least-squares linear models. Pooled CBS-control models adjusted for diagnostic group, age at encounter, and education, whereas CBS-only models adjusted for age at encounter, duration from symptom onset to examination, and education. Candidate predictor models for sensorimotor DTI-MD were treated as targeted exploratory analyses and were summarized using adjusted beta estimates, 95% confidence intervals, p-values, and adjusted R² rather than FDR-corrected inference. Amyloid-positive versus amyloid-negative CBS sensitivity analyses used ROI-wise Welch two-sample t-tests with Benjamini-Hochberg FDR correction across tested ROIs. All statistical tests were two-sided. Given the exploratory nature of this work, results emphasize effect sizes, model estimates, confidence intervals, and anatomical patterns, with findings interpreted as hypothesis-generating.

## Results

### Cohort characteristics

The study included 32 participants with clinically diagnosed CBS. Participants had a median age at visit of 68 years and a median disease duration of 3 years from symptom onset. **Table 1** shows demographics did not differ between CBS and control groups, with no differences in age, education, sex distribution, or *APOE* ε4 carrier status. Compared with controls, the CBS group demonstrated worse performance on MoCA, MDS-UPDRS III, WAB-Praxis and TULIA. In detail, the cognitive impairment was mild to moderate (median MoCA score of 22), with relatively preserved executive function (median FAB score of 15). Motor severity was moderate, as reflected by MDS-UPDRS III and PSP Rating Scale scores. Consistent with the clinical phenotype of CBS, participants demonstrated prominent praxis impairment and asymmetric limb involvement, as captured by WAB-Praxis and TULIA subscores.

**Table 1.** Demographic and clinical characteristics of study participants. Data is median (IQR) for continuous variables and n (%) for categorical variables. Group differences were assessed using the Kruskal-Wallis test (continuous) and chi-square or Fisher’s exact test (categorical). Clinical scales: MoCA (Montreal Cognitive Assessment); FAB (Frontal Assessment Battery); MDS-UPDRS III (Movement Disorder Society-sponsored revision of the Unified Parkinson’s Disease Rating Scale, Part III); PSP Rating Scale (Progressive Supranuclear Palsy Rating Scale); PSIS (PSP Saccadic Impairment Scale); WAB-Praxis (Western Aphasia Battery - Praxis Subtest); TULIA total (Test of Upper Limb Apraxia); TULIA-Left and TULIA-Right (left/right subscores). FAB and PSP Rating Scale were collected in CBS participants as part of the clinical research assessment but were not collected in controls; group comparisons were therefore not performed. Statistical significance was set at p < 0.05.

| Variable | Control (n=35) | CBS (n=32) | P across all groups |
| --- | --- | --- | --- |
| Demographic and disease characteristics |  |  |  |
| Age at visit, years | 66 (63, 70) | 68 (65, 72) | 0.16 |
| Age at onset, years | N/A | 65 (60, 68) | N/A |
| Disease duration, years | N/A | 3 (2, 4) | N/A |
| Education, years | 16 (15, 16) | 14 (14, 16) | 0.08 |
| Categorical clinical features |  |  |  |
| Female, N (%) | 18 (51%) | 17 (53%) | 0.89 |
| Non-right-handed, N (%) | 3 (9%) | 3 (9%) | 0.99 |
| Family history, N (%) | 0 (0%) | 8 (25%) | 0.002 |
| APOE ε4 carrier | 4 (11%) | 8 (25%) | 0.15 |
| Apraxia of speech present | 0 (0%) | 7 (22%) | 0.004 |
| Aphasia present | 0 (0%) | 4 (12%) | 0.047 |
| Neurological measures |  |  |  |
| MoCA (/ 30) | 27 (26, 28) | 22 (20, 24) | <0.001 |
| MDS-UPDRS III (/ 132) | 1 (0, 2) | 32 (20, 47) | <0.001 |
| PSP Rating Scale (/100) | N/A | 25 (21, 36) | N/A |
| FAB (/18) | N/A | 15 (12, 17) | N/A |
| WAB Praxis (/60) | 60 (58, 60) | 52 (49, 55) | <0.001 |
| PSIS (/5) | 0 (0, 0) | 1 (0, 1) | <0.001 |
| TULIA (/12) | 12 (11, 12) | 5 (2, 9) | <0.001 |
| TULIA (left) (/12) | 12 (11, 12) | 9 (7, 10) | <0.001 |
| TULIA (right) (/12) | 12 (11, 12) | 10 (5, 11) | 0.01 |

### Regional measures best separate CBS from control

We first examined the extent and distribution of ROI-level CBS-control differences across modalities (**Figure 2**). dMRI abnormalities were widespread across WM ROIs (**Table S2**), whereas tau PET abnormalities were comparatively sparse. DTI MD showed the broadest CBS-control separation, with 86/88 (98%) ROIs significant after FDR correction. NODDI ICVF also showed widespread abnormalities, with 67/88 (76%) ROIs FDR-significant. Structural volume showed more focal patterns than dMRI, with FDR-significant abnormalities in 47/120 (39%) ROIs. The original tau PET images showed few nominal ROI-level differences and no FDR-significant ROIs. PC1-removed tau showed more ROI-level separation than original tau but remained less widespread than dMRI and structural MRI. The ROIs contributing to the figures are shown in **Table S1** and **S2**.

**Figure 2.**
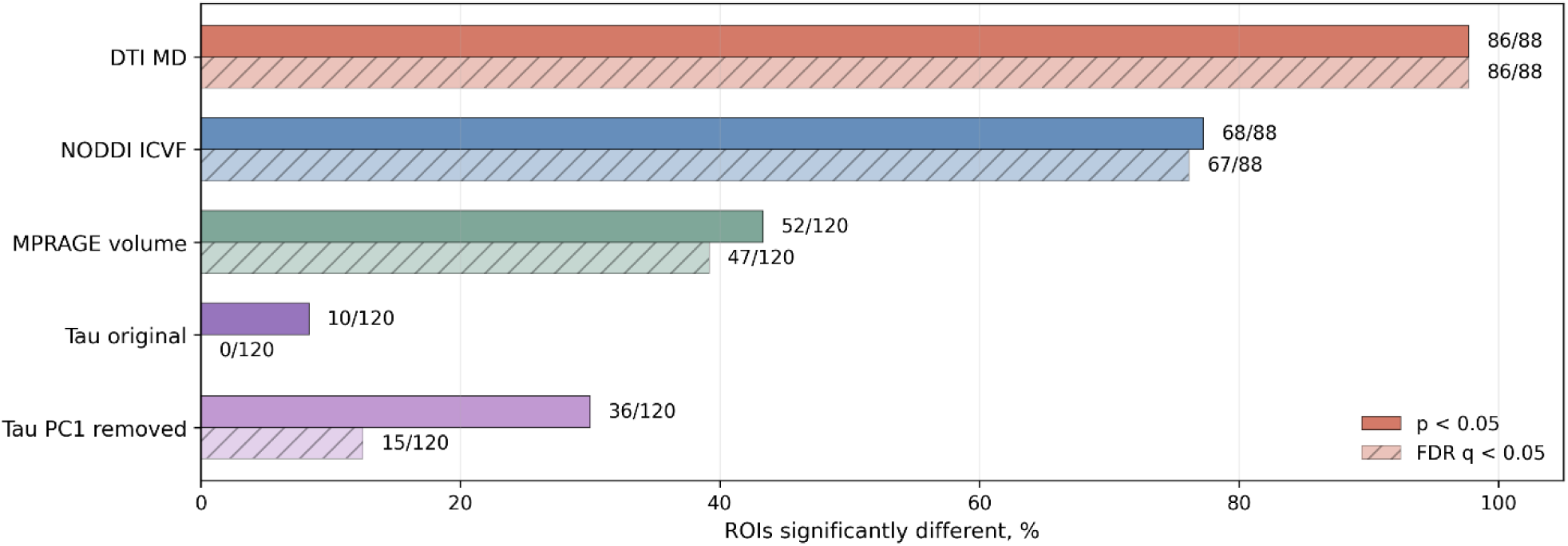
Extent and distribution of CBS-control imaging abnormalities across modalities. Percentage of ROIs showing CBS-control group differences at nominal p<0.05 and after FDR correction within each modality. dMRI measures showed widespread ROI-level abnormalities across JHU EVE WM ROIs, particularly DTI MD and NODDI ICVF, whereas tau PET showed fewer ROI-level group differences.

When distinguishing CBS from controls, the strongest and most consistent signal came from dMRI, with the greatest AUCs observed for MD- and ICVF-based measures, particularly in precentral and postcentral WM, superior corona radiata and the posterior limb of the internal capsule (**Figure 3A and Figure S1)**. MD/ICVF in precentral WM showed high AUC ∼0.95 (MD: Cohen’s d = 2.2, p value ∼ E-10, ICVF: Cohen’s d = −2.4, p value ∼ E-10, **Table S2**). The next highest AUCs were obtained with volume, with the greatest differentiation between CBS and controls obtained with precentral and basal ganglia regions (**Figure S1**). Tau PET showed the smallest AUC values, with group separation effects higher for PC1-subtracted compared to unadjusted tau PET. Regions that showed the best group separation for PC1-subtracted tau PET included pallidum and precentral gyrus (**Figure S1**). Comparative analyses of both AUC and absolute Cohen’s *d* demonstrated the relative ability of each modality to distinguish CBS from controls, ranked as follows: MD and ICVF > other dMRI metrics > structural volume > tau PET with PC1-subtracted ≫ unadjusted tau PET (**Figure 3B–C**). The ROIs contributing to the figures are attached to Table S1 and S2. The data supporting Figure 3 is shown in **Table S4**.

**Figure 3.**
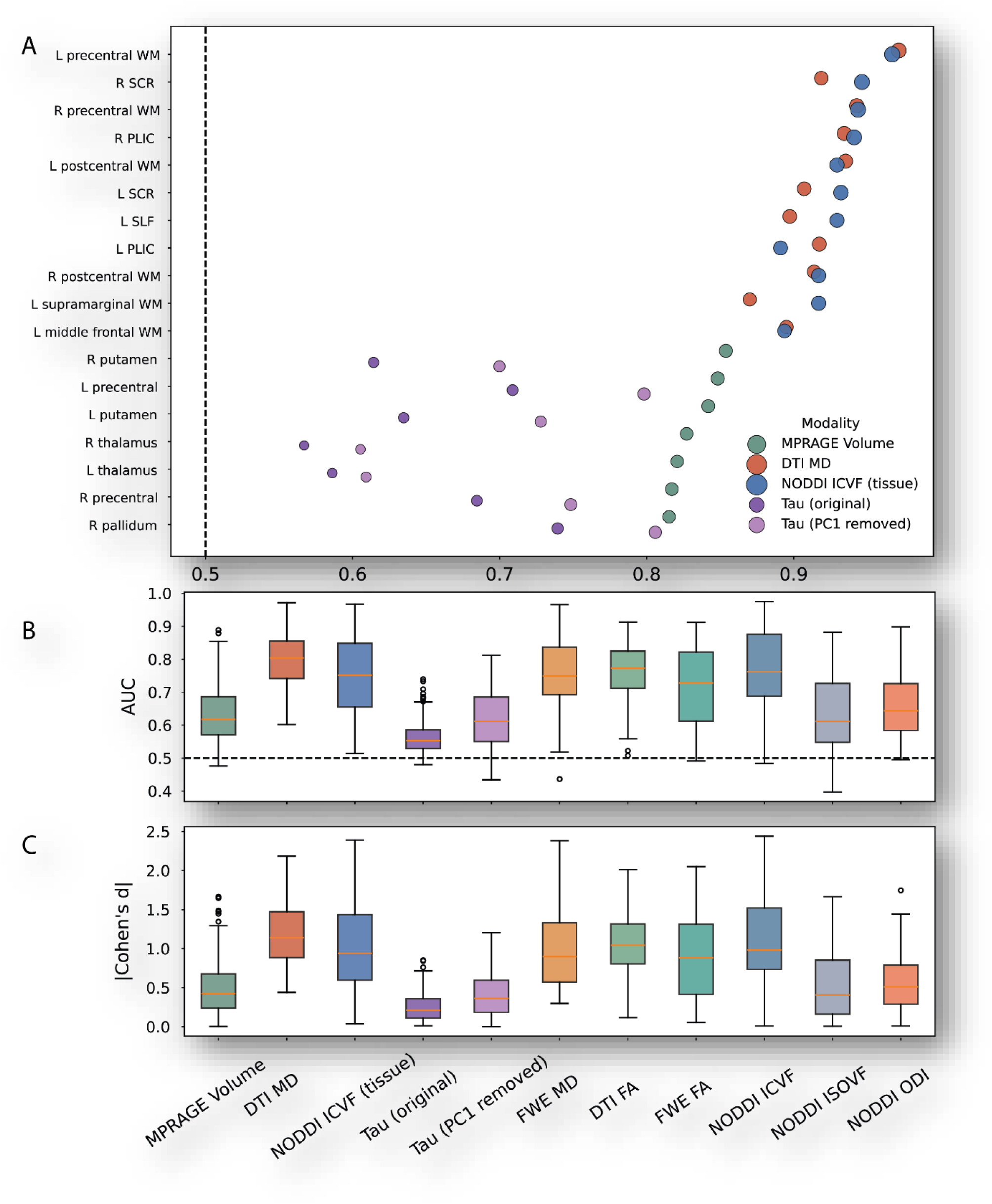
Top discriminative imaging features across metrics and ROIs. **(A)** Univariate AUC for distinguishing CBS from controls using DTI MD, NODDI ICVF, structural volume, tau PET with the first principal component removed, and unadjusted tau PET. ROIs shown in Fig. 2A were first ranked within each modality, and the final displayed set was restricted to shared ROIs with the highest overall cross-modality AUC for visual comparability and space constraints. Modality-specific top ROIs are shown in Fig. S1. **(B)** AUC-based ranking of all metrics. **(C)** Ranking of metrics by absolute effect size (Cohen’s *d*). Abbreviations: CBS, corticobasal syndrome; AUC, area under the receiver operating characteristic curve; DTI, diffusion tensor imaging; MD, mean diffusivity; FA, fractional anisotropy; FWE, free-water-eliminated; NODDI, neurite orientation dispersion and density imaging; ICVF, intracellular volume fraction; ISOVF, isotropic volume fraction; ODI, orientation dispersion index; PET, positron emission tomography; ROI, region of interest; PC1, first principal component.

Bootstrap comparison of sensorimotor summary AUCs supported the stronger CBS-control discrimination of dMRI metrics: DTI MD (AUC 0.976, 95% bootstrap CI 0.934-1.000) and NODDI ICVF (AUC 0.935, CI 0.864-0.988) showed higher AUCs than structural volume (GM AUC 0.618, CI 0.477-0.754, WM AUC 0.767, CI 0.642-0.880) and tau PET measures (Tau PC1 removed AUC 0.688, CI 0.540-0.825, Tau AUC 0.622, CI 0.467-0.768), with all dMRI versus non-dMRI comparisons remaining significant after FDR correction (**Table S5**).

### Sensorimotor dMRI relationships to volume and tau

Correlation analyses demonstrated that lower sensorimotor GM volume was associated with greater sensorimotor WM MD in both pooled CBS and control (R²=0.73, p=6.2×10^−5^) (**Figure 4A**) and CBS-only (R²=0.53, p=1.5×10^−4^) models (**Figure 4B)**. Sensorimotor WM volume showed a similar but weaker association in the pooled sample (R²=0.66, p=0.059) (**Figure 4C)** but not in the CBS-only model (R²=0.25, p=0.157) (**Figure 4D)**.

**Figure 4.**
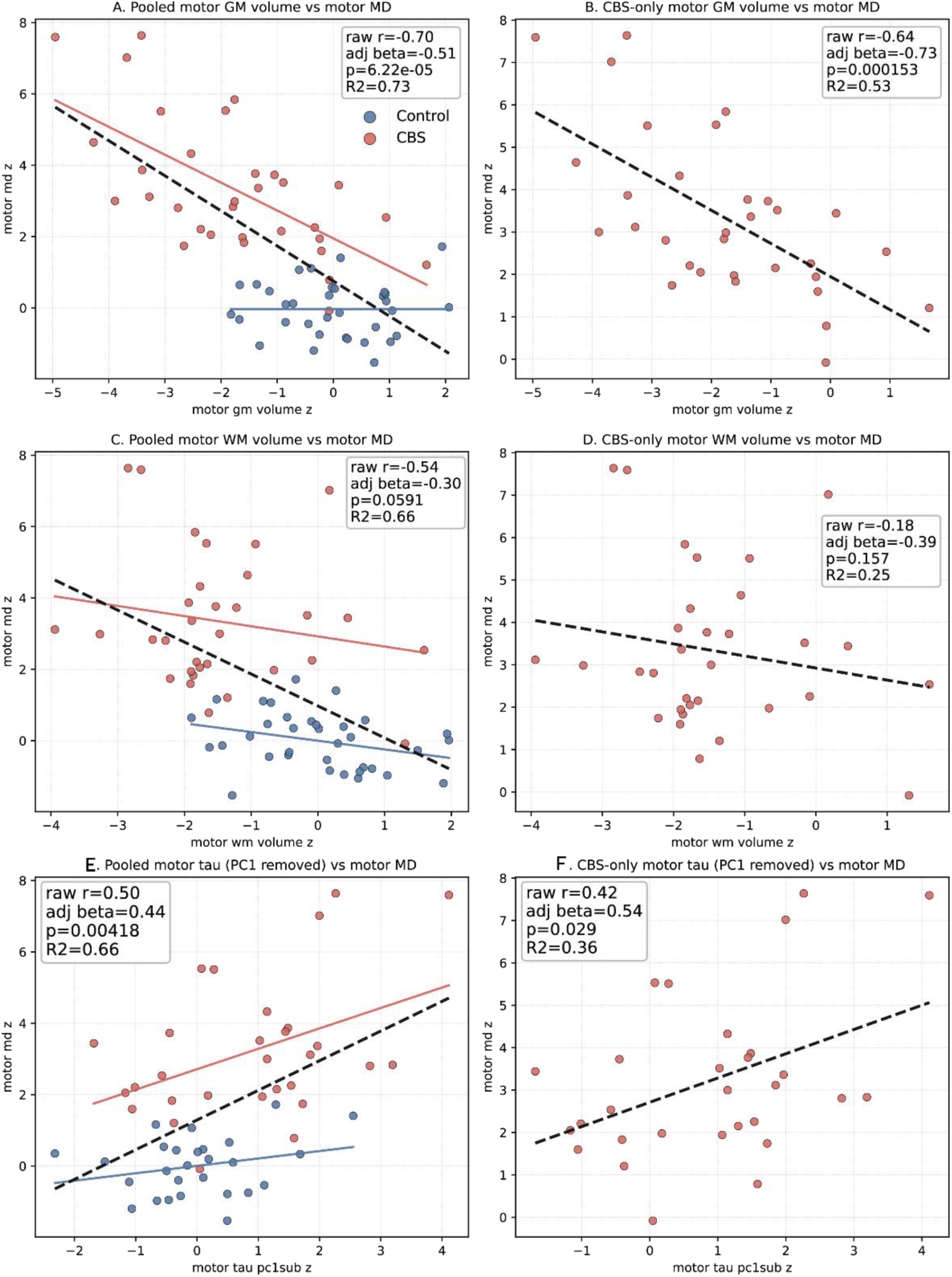
Associations of sensorimotor GM volume z-score and tau PET uptake with sensorimotor WM microstructural abnormality. Panels A-B show the relationship between sensorimotor GM volume and sensorimotor WM DTI-MD z-score in the pooled CBS and control sample (A) and in CBS participants only (B). Panels C-D show the relationship between sensorimotor WM volume and sensorimotor WM DTI MD z-score in the pooled sample (C) and in CBS participants only (D). Panels E-F show the relationship between PC1-removed sensorimotor tau PET z-score and sensorimotor WM DTI MD z-score in the pooled sample (E) and in CBS participants only (F). Sensorimotor GM volume and tau PET measures were summarized across precentral, postcentral, Rolandic opercular, supplementary motor, and paracentral regions. Sensorimotor WM volume and DTI MD were summarized across precentral WM, postcentral WM, and corticospinal tract ROIs. Lines show fitted linear relationships; pooled panels include both CBS and control participants, whereas CBS-only panels test within-disease variability.

Tau uptake after PC1-subtraction (**Figure 4E, F)** showed higher sensorimotor tau uptake related to greater sensorimotor MD in the pooled sample (R²=0.66, p=0.004) and in CBS alone (R²=0.36, p=0.029).

### Candidate correlates of sensorimotor diffusion abnormalities

To compare candidate correlates of sensorimotor diffusion abnormalities, we fit CBS-only single-predictor models relating volume and tau measures to sensorimotor DTI-MD, adjusting for age at encounter, symptom duration, and education. **Figure 5A** ranks each predictor by adjusted R², and **Figure 5B** shows the corresponding adjusted beta estimates. Greater sensorimotor GM volume loss showed the strongest association with higher sensorimotor DTI-MD (β=0.73, 95% CI 0.39 to 1.07, p=0.00015; adjusted R²=0.46). Higher PC1-removed sensorimotor tau uptake was also associated with higher sensorimotor DTI-MD (β=0.54, 95% CI 0.06 to 1.02, p=0.029; adjusted R²=0.25). In contrast, original sensorimotor tau uptake, subcortical tau uptake with or without PC1 removal, subcortical GM volume loss, and cortical GM volume loss were not significantly associated with sensorimotor DTI-MD. Thus, among the tested candidate predictors, sensorimotor GM volume loss showed the strongest relationship with sensorimotor DTI-MD, with an additional association observed for PC1-removed sensorimotor tau.

**Figure 5.**
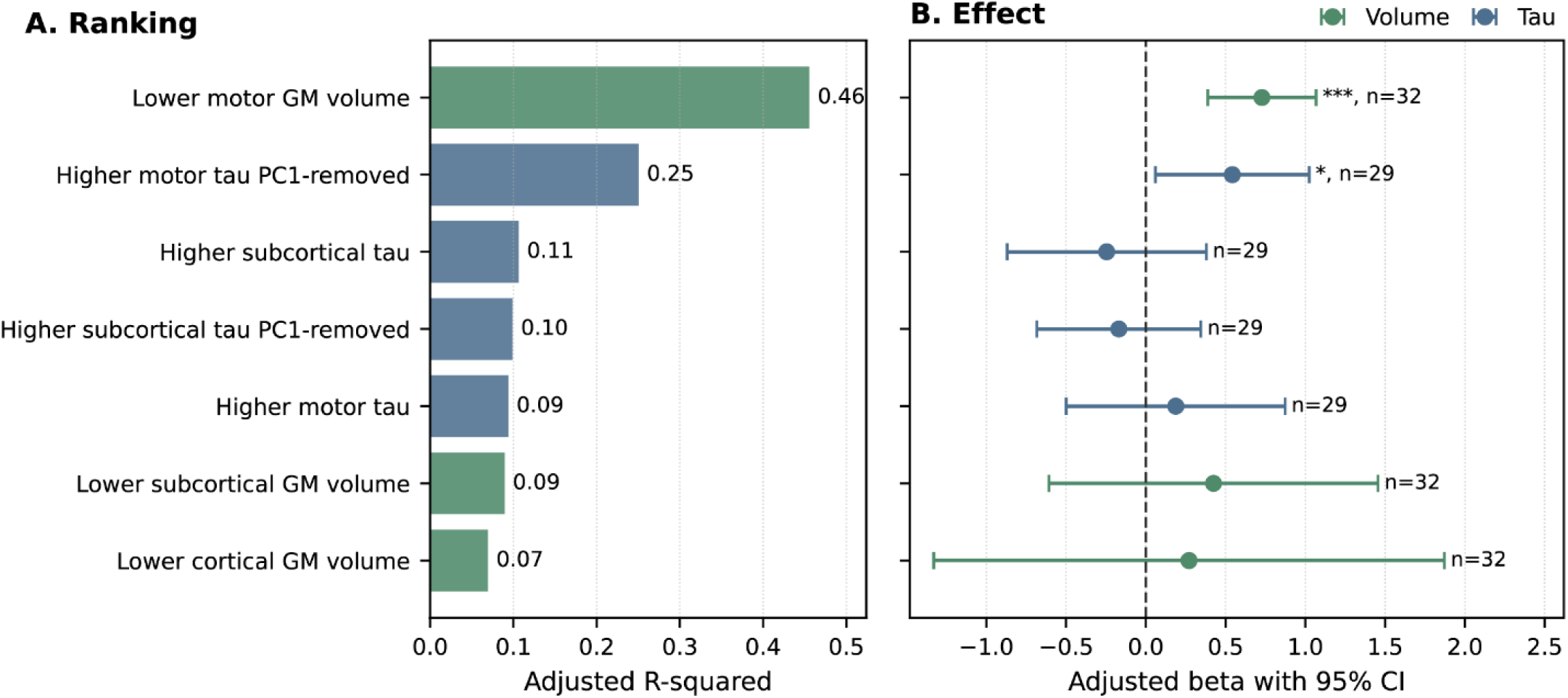
Candidate predictors of sensorimotor DTI-MD in CBS. CBS-only single-predictor Ordinary Least Squares models tested whether sensorimotor DTI-MD was associated with GM volume loss or tau uptake after adjustment for age at encounter, symptom duration, and education. Panel A ranks predictors by adjusted R². Panel B shows covariate-adjusted beta coefficients with 95% confidence intervals; the dashed vertical line indicates β=0. Lower GM volume indicates greater GM volume loss, coded as the negative of the control-normalized volume z-score. Higher tau indicates greater control-normalized tau uptake; PC1-removed tau denotes tau uptake after removal of the first principal component signal. Subcortical composites included bilateral caudate, pallidum, and thalamus. *p<0.05, **p<0.01, ***p<0. 001.

### Amyloid Status Sensitivity Analysis

Because a small number of CBS participants were amyloid-positive, we examined whether amyloid status influenced regional imaging measures within the CBS group. Among CBS participants with available amyloid PET, 4 were amyloid-positive (15%) and 23 were amyloid-negative (85%); 5 participants without amyloid PET were excluded from this sensitivity analysis. ROI-wise values were compared between the amyloid-positive and amyloid-negative CBS participants across DTI-MD, tissue-corrected NODDI ICVF, original tau PET, and PC1-removed tau PET measures. No differences survived FDR correction (**Table S6**).

Controls were not selected based on amyloid status; among controls with available PiB PET, 8/30 were amyloid-positive and 22/30 were amyloid-negative. Because the amyloid-status sensitivity analysis was performed within the CBS group, control amyloid status did not enter that analysis. In a post hoc check restricting the control group to amyloid-negative controls, the relative pattern of sensorimotor discrimination remained consistent with the primary AUC analysis in **Figure 3/Table S5**, with dMRI measures retaining higher AUCs (MD 0.977) than tau PET measures (PC1-removed tau 0.659).

## Discussion

This multimodal study showed that CBS is associated with widespread and severe disruptions to WM integrity measured using dMRI, with dMRI providing better differentiation of CBS from controls than either structural MRI or tau PET. Abnormalities on dMRI were related both to GM volume loss and tau PET uptake within the sensorimotor cortex, although GM volume was the strongest predictor of dMRI abnormalities. Hence, dMRI, particularly DTI-MD and NODDI-ICVF, provides the most promising diagnostic biomarkers for CBS.

Across modalities, dMRI provided the most extensive discrimination between CBS and controls, particularly for MD and ICVF measures. DTI-MD abnormalities were widespread, with the strongest effects in sensorimotor and projection WM pathways, including precentral and postcentral WM, posterior limb of the internal capsule, superior corona radiata, superior longitudinal fasciculus, and corpus callosum. Tissue-corrected NODDI ICVF showed a complementary pattern, with lower ICVF in many of the same motor and projection pathways. This regional distribution is consistent with prior CBS and atypical parkinsonism studies showing prominent involvement of corticospinal, callosal, corona radiata, and perirolandic WM pathways(Tian, Ali et al. 2026). The prominence of MD and ICVF is biologically plausible: MD is sensitive to broad microstructural disorganization, gliosis, and tissue rarefaction, whereas ICVF is more specifically indexes loss or simplification of neurite architecture. Together, these metrics may capture downstream WM injury more sensitively than macrostructural volume or first-generation tau PET measures.

Structural volume did not perform as well as dMRI but still showed excellent differentiation of CBS and controls. The greatest volume loss was observed in the sensorimotor cortex and subcortical structures, including precentral cortex, putamen, thalamus, and pallidum, consistent with the known motor cortical and basal ganglia involvement of CBS(Ali, Whitwell et al. 2018) (Nakano, Shimada et al. 2022). Although both GM and WM volume measures were examined, GM volume provided a stronger relationship with sensorimotor DTI-MD. This suggests that WM diffusion abnormalities may partly reflect degeneration linked to Wallerian degeneration, rather than simply local WM volume loss. The finding that sensorimotor GM volume was the strongest predictor of sensorimotor DTI-MD supports a network-level interpretation in which cortical motor-system degeneration and downstream WM microstructural injury evolve together.

Tau PET showed weaker and less spatially extensive CBS-control differentiation than dMRI or volume. Original flortaucipir uptake showed limited group separation and weak associations with sensorimotor volume and dMRI measures. This pattern is consistent with prior CBS flortaucipir studies showing marked heterogeneity and even minimal uptake in clinically typical cases; notably, (Ali, Whitwell et al. 2018) reported no elevated AV-1451 uptake in PiB-negative typical CBS without apraxia of speech (AOS), whereas elevated uptake was more apparent in AOS-associated CBS phenotypes(Smith, Schöll et al. 2017). After PC1 removal, PET findings became more interpretable, with stronger signals in regions including pallidum and precentral cortex and a significant association between higher PC1-removed sensorimotor tau and higher sensorimotor DTI-MD. However, this association was weaker than the volume-DTI-MD relationship, and sensorimotor GM volume remained the strongest predictor in the candidate-predictor models. These results suggest that PC1 removal may reduce a dominant global or nonspecific component that obscures clinically relevant regional tau signal, as previously demonstrated in progressive supranuclear palsy(Satoh, Ali et al. 2026), but also that flortaucipir PET captures only part of the structural injury reflected by dMRI.

However, the tau PET findings should be interpreted cautiously considering known limitations of first-generation flortaucipir PET in non-Alzheimer 4R tauopathies. Autoradiography studies show that flortaucipir binds strongly to AD-type paired-helical filament tau but has relatively low or inconsistent binding in PSP/CBD and other non-AD tauopathies, with additional off-target signal that can complicate regional interpretation (Lowe, Curran et al. 2016) (Sander, Lashley et al. 2016) (Aguero, Dhaynaut et al. 2019). Because CBS is pathologically heterogeneous and often reflects corticobasal degeneration or PSP, these properties likely reduce the sensitivity and regional specificity of flortaucipir in this syndrome. Thus, the relative underperformance of tau PET compared with dMRI does not necessarily imply that tau pathology is unimportant in CBS; rather, it may reflect limitations of the tracer and the complex relationship between molecular pathology and downstream tissue injury.

A major strength of this study is the direct comparison of dMRI, structural volume, and tau PET within the same clinically well-characterized CBS cohort, with all participants undergoing a standardized neurologic and imaging evaluation. This design reduces between-cohort variability and strengthens inferences regarding the relative sensitivity of each modality. Limitations include the use of first-generation flortaucipir, which has known limitations in non-Alzheimer 4R tauopathies, and the modest sample size, which reduces power for subgroup analyses and for detecting subtler regional effects. The CBS cohort in this study was selected to remove individuals with biomarker confirmed AD, i.e. those with both global amyloid and temporal lobe tau uptake suggestive of AD. However, four patients that remained in the cohort showed evidence of amyloid deposition on PET. Although the research criteria suggest only the use of amyloid-PET to exclude AD, the addition of tau PET increases specificity. Furthermore, the criteria was published prior to the development of tau PET. We did not find any evidence of imaging differences between these four patients and the rest of the cohort, but interpretation is limited by the small amyloid-positive subgroup.

Overall, these findings support dMRI as the most sensitive imaging measure of regional CBS-related tissue injury. The strongest abnormalities were localized to sensorimotor and projection WM pathways, while volume and PC1-removed tau measures provided complementary but less extensive information. A practical implication is that dMRI, especially MD and ICVF, may be particularly useful for detecting and quantifying disease-related network injury in CBS and to serve as supportive diagnostic biomarkers. Tau PET may require improved tracers or signal-processing approaches to better capture non-Alzheimer tau pathology in CBS.

## Acknowledgements

This study was funded by National Institutes of Health (NIH) grants R01-AG87140, R01-DC12519 and R01-NS89757.

## Data Availability

The data supporting the findings of this study are available from the corresponding author upon reasonable request.

