## Supplemental for "Diffusion MRI Is More Sensitive Than Tau PET and Regional Volume Loss in Corticobasal Syndrome"

**Supplemental materials**

**ROI definition for multiple modalities**

MPRAGE GM volume and tau PET analyses used ADIR122/MCALT atlas regions, with left and right homologous regions analyzed separately at the subject level. MPRAGE values were TIV-adjusted regional tissue volumes, whereas tau PET values were regional mean uptake values for original tau and PC1-removed tau. dMRI analyses used JHU TypeIII white-matter ROIs and z-score metrics. Dorsal mesopontine and pons were excluded from the regenerated modality-comparison summary. The tau source data included an Unknown atlas label, which is retained for reproducibility.

DMRI and MPRAGE WM volume analyses were performed using JHU TypeIII white-matter ROIs. Left and right homologous tracts were analyzed separately at the subject level. dMRI metrics included DTI MD, NODDI ICVF (tissue), FWE MD, DTI FA, FWE FA, NODDI ICVF, NODDI ISOVF, and NODDI ODI, each expressed as control-referenced z-scores.

**Table S1. GM ROIs included in the modality-comparison analyses**.

| Roi_name_clean | hemisphere |
|---|---|
| **Amygdala left** | Left |
| **Amygdala right** | Right |
| **Angular left** | Left |
| **Angular right** | Right |
| **Calcarine left** | Left |
| **Calcarine right** | Right |
| **Caudate left** | Left |
| **Caudate right** | Right |
| **Cerebellum 10 left** | Left |
| **Cerebellum 10 right** | Right |
| **Cerebellum 3 left** | Left |
| **Cerebellum 3 right** | Right |
| **Cerebellum 4 5 left** | Left |
| **Cerebellum 4 5 right** | Right |
| **Cerebellum 6 left** | Left |
| **Cerebellum 6 right** | Right |
| **Cerebellum 7b left** | Left |
| **Cerebellum 7b right** | Right |
| **Cerebellum 8 left** | Left |
| **Cerebellum 8 right** | Right |
| **Cerebellum 9 left** | Left |
| **Cerebellum 9 right** | Right |
| **Cerebellum Crus1 left** | Left |
| **Cerebellum Crus1 right** | Right |
| **Cerebellum Crus2 left** | Left |
| **Cerebellum Crus2 right** | Right |
| **Cingulum Ant left** | Left |
| **Cingulum Ant right** | Right |
| **Cingulum Mid left** | Left |
| **Cingulum Mid right** | Right |
| **Cingulum Post left** | Left |

| | |
|---|---|
| **Cingulum Post right** | Right |
| **Cuneus left** | Left |
| **Cuneus right** | Right |
| **Entorhinal Cortex left** | Left |
| **Entorhinal Cortex right** | Right |
| **Frontal Inf Oper left** | Left |
| **Frontal Inf Oper right** | Right |
| **Frontal Inf Orb left** | Left |
| **Frontal Inf Orb right** | Right |
| **Frontal Inf Tri left** | Left |
| **Frontal Inf Tri right** | Right |
| **Frontal Med Orb left** | Left |
| **Frontal Med Orb right** | Right |
| **Frontal Mid left** | Left |
| **Frontal Mid Orb left** | Left |
| **Frontal Mid Orb right** | Right |
| **Frontal Mid right** | Right |
| **Frontal Sup left** | Left |
| **Frontal Sup Medial left** | Left |
| **Frontal Sup Medial right** | Right |
| **Frontal Sup Orb left** | Left |
| **Frontal Sup Orb right** | Right |
| **Frontal Sup right** | Right |
| **Fusiform left** | Left |
| **Fusiform right** | Right |
| **Heschl left** | Left |
| **Heschl right** | Right |
| **Hippocampus left** | Left |
| **Hippocampus right** | Right |
| **Insula left** | Left |
| **Insula right** | Right |
| **Lingual left** | Left |
| **Lingual right** | Right |
| **Occipital Inf left** | Left |
| **Occipital Inf right** | Right |
| **Occipital Mid left** | Left |
| **Occipital Mid right** | Right |
| **Occipital Sup left** | Left |
| **Occipital Sup right** | Right |
| **Olfactory left** | Left |
| **Olfactory right** | Right |
| **Pallidum left** | Left |
| **Pallidum right** | Right |
| **Paracentral Lobule left** | Left |
| **Paracentral Lobule right** | Right |
| **ParaHippocampal left** | Left |
| **ParaHippocampal right** | Right |
| **Parietal Inf left** | Left |
| **Parietal Inf right** | Right |
| **Parietal Sup left** | Left |
| **Parietal Sup right** | Right |

| | |
|---|---|
| **Postcentral left** | Left |
| **Postcentral right** | Right |
| **Precentral left** | Left |
| **Precentral right** | Right |
| **Precuneus left** | Left |
| **Precuneus right** | Right |
| **Putamen left** | Left |
| **Putamen right** | Right |
| **Rectus left** | Left |
| **Rectus right** | Right |
| **Retrosplenial Cortex left** | Left |
| **Retrosplenial Cortex right** | Right |
| **Rolandic Oper left** | Left |
| **Rolandic Oper right** | Right |
| **Supp Motor Area left** | Left |
| **Supp Motor Area right** | Right |
| **SupraMarginal left** | Left |
| **SupraMarginal right** | Right |
| **Temporal Inf left** | Left |
| **Temporal Inf right** | Right |
| **Temporal Mid left** | Left |
| **Temporal Mid right** | Right |
| **Temporal Pole Mid left** | Left |
| **Temporal Pole Mid right** | Right |
| **Temporal Pole Sup left** | Left |
| **Temporal Pole Sup right** | Right |
| **Temporal Sup left** | Left |
| **Temporal Sup right** | Right |
| **Thalamus left** | Left |
| **Thalamus right** | Right |
| **Vermis 1 2** | Midline/Unspecified |
| **Vermis 10** | Midline/Unspecified |
| **Vermis 3** | Midline/Unspecified |
| **Vermis 4 5** | Midline/Unspecified |
| **Vermis 6** | Midline/Unspecified |
| **Vermis 7** | Midline/Unspecified |
| **Vermis 8** | Midline/Unspecified |
| **Vermis 9** | Midline/Unspecified |

**Table S2 WM ROIs from JHU atlas.**

| roi_name_clean | hemisphere |
|---|---|
| Angular WM Left (AWM L) | Left |
| Angular WM Right (AWM R) | Right |
| Anterior Corona Radiata Left (ACR L) | Left |
| Anterior Corona Radiata Right (ACR R) | Right |
| Anterior Limb of Internal Capsule Left (ALIC L) | Left |
| Anterior Limb of Internal Capsule Right (ALIC R) | Right |
| Body of Corpus Callosum Left (BCC L) | Left |
| Body of Corpus Callosum Right (BCC R) | Right |
| Cerebral Peduncle Left (CP L) | Left |
| Cerebral Peduncle Right (CP R) | Right |
| Cingulum (cingulate Gyrus) Left (CGC L) | Left |
| Cingulum (cingulate Gyrus) Right (CGC R) | Right |
| Cingulum (hippocampus) Left (CGH L) | Left |
| Cingulum (hippocampus) Right (CGH R) | Right |
| Corticospinal Tract Left (CST L) | Left |
| Corticospinal Tract Right (CST R) | Right |
| External Capsule Left (EC L) | Left |
| External Capsule Right (EC R) | Right |
| Fornix (column and Body) Left (Fx L) | Left |
| Fornix (column and Body) Right (Fx R) | Right |
| Fornix(cres) Stria Terminalis Right (Fx ST R) | Right |
| Fornix(cres) Stria Terminalisleft (Fx ST L) | Left |
| Frontal orbital | Midline/Unspecified |
| Fusiform WM Left (FuWM L) | Left |
| Fusiform WM Right (FuWM R) | Right |
| Genu of Corpus Callosum Left (GCC L) | Left |

| | |
|---|---|
| **Genu of Corpus Callosum Right (GCC R)** | Right |
| **Inferior Cerebellar Peduncle Left (ICP L)** | Left |
| **Inferior Cerebellar Peduncle Right (ICP R)** | Right |
| **Inferior Frontal WM Left (IFWM L)** | Left |
| **Inferior Frontal WM Right (IFWM R)** | Right |
| **Inferior Fronto-occipital Fasciculus Left (IFO L)** | Left |
| **Inferior Fronto-occipital Fasciculus Right (IFO R)** | Right |
| **Inferior Temporal WM Left (ITWM L)** | Left |
| **Inferior Temporal WM Right (ITWM R)** | Right |
| **Medial Lemniscus Left (ML L)** | Left |
| **Medial Lemniscus Right (ML R)** | Right |
| **Midbrain Left (Midbrain L)** | Left |
| **Midbrain Right (Midbrain R)** | Right |
| **Middle Cerebellar Peduncle Left (MCP L)** | Left |
| **Middle Cerebellar Peduncle Right (MCP R)** | Right |
| **Middle Frontal WM Left (MFWM L)** | Left |
| **Middle Frontal WM Right (MFWM R)** | Right |
| **Middle Temporal WM Left (MTWM L)** | Left |
| **Middle Temporal WM Right (MTWM R)** | Right |
| **Occipital lateral** | Midline/Unspecified |
| **Occipital medial** | Midline/Unspecified |
| **Pontine Crossing Tract (a Part of MCP) Right (PCT R)** | Right |
| **Pontine Crossing Tract Left (PCT L)** | Left |
| **Postcentral WM Left (PoCWM L)** | Left |
| **Postcentral WM Right (PoCWM R)** | Right |
| **Posterior Corona Radiata Left (PCR L)** | Left |
| **Posterior Corona Radiata Right (PCR R)** | Right |

| | |
|---|---|
| **Posterior Limb of Internal Capsule Left (PLIC L)** | Left |
| **Posterior Limb of Internal Capsule Right (PLIC R)** | Right |
| **Posterior Thalamic Radiation Left (PTR L)** | Left |
| **Posterior Thalamic Radiation Right (PTR R)** | Right |
| **Pre-cuneus WM Left (PreCuWM L)** | Left |
| **Pre-cuneus WM Right (PreCuWM R)** | Right |
| **Precentral WM Left (PrCWM L)** | Left |
| **Precentral WM Right (PrCWM R)** | Right |
| **Retrolenticular Part of Internal Capsule Left (RLIC L)** | Left |
| **Retrolenticular Part of Internal Capsule Right (RLIC R)** | Right |
| **Sagittal Stratum Left (SS L)** | Left |
| **Sagittal Stratum Right (SS R)** | Right |
| **Splenium of Corpus Callosum Left (SCC L)** | Left |
| **Splenium of Corpus Callosum Right (SCC R)** | Right |
| **Superior Cerebellar Peduncle Left (SCP L)** | Left |
| **Superior Cerebellar Peduncle Right (SCP R)** | Right |
| **Superior Corona Radiata Left (SCR L)** | Left |
| **Superior Corona Radiata Right (SCR R)** | Right |
| **Superior Frontal WM Left (SFWM L)** | Left |
| **Superior Frontal WM Right (SFWM R)** | Right |
| **Superior Fronto-occipital Fasciculus Left (SFO L)** | Left |
| **Superior Fronto-occipital Fasciculus Right (SFO R)** | Right |
| **Superior Longitudinal Fasciculus Left (SLF L)** | Left |
| **Superior Longitudinal Fasciculus Right (SLF R)** | Right |
| **Superior Parietal WM Left (SPWM L)** | Left |

| | |
|---|---|
| **Superior Parietal WM Right (SPWM R)** | Right |
| **Superior Temporal WM Left (STWM L)** | Left |
| **Superior Temporal WM Right (STWM R)** | Right |
| **Supramarginal WM Left (SMWM L)** | Left |
| **Supramarginal WM Right (SMWM R)** | Right |
| **Tapatum Left (TAP L)** | Left |
| **Tapatum Right (TAP R)** | Right |
| **Temporal medial** | Midline/Unspecified |
| **Uncinate Fasciculus Left (UNC L)** | Left |
| **Uncinate Fasciculus Right (UNC R)** | Right |

**Table S3 Sensorimotor GM and WM ROIs**

| MODALITY | ROI |
|---|---|
| **GM VOLUME** | Postcentral_L |
| **GM VOLUME** | Postcentral_R |
| **GM VOLUME** | Precentral_L |
| **GM VOLUME** | Precentral_R |
| **GM VOLUME** | Rolandic_Oper_L |
| **GM VOLUME** | Rolandic_Oper_R |
| **GM VOLUME** | Supp_Motor_Area_L |
| **GM VOLUME** | Supp_Motor_Area_R |
| **GM VOLUME** | Paracentral_Lobule_L |
| **GM VOLUME** | Paracentral_Lobule_R |
| **WM VOLUME** | CST_L Corticospinal_tract_left |
| **WM VOLUME** | CST_R Corticospinal_tract_right |
| **WM VOLUME** | PoCWM_L POSTCENTRAL__WM_left |
| **WM VOLUME** | PoCWM_R POSTCENTRAL_WM_right |
| **WM VOLUME** | PrCWM_L PRECENTRAL__WM_left |
| **WM VOLUME** | PrCWM_R PRECENTRAL_WM_right |

**Top ROI predictors across modalities in the combined comparison**

**Figure S1 Top ROI predictors across modalities in the combined MPRAGE-Tau comparison.** Dots show the top-ranked ROIs for MPRAGE volume, DTI MD, NODDI ICVF, original tau, and PC1-removed tau, ranked by univariate AUC for distinguishing CBS from controls. Dot size scales with absolute Cohen's d, and color indicates modality. The vertical dashed line marks AUC = 0.5. Original tau and PC1-removed tau are shown separately to illustrate the effect of PC1 removal on tau-based discrimination.

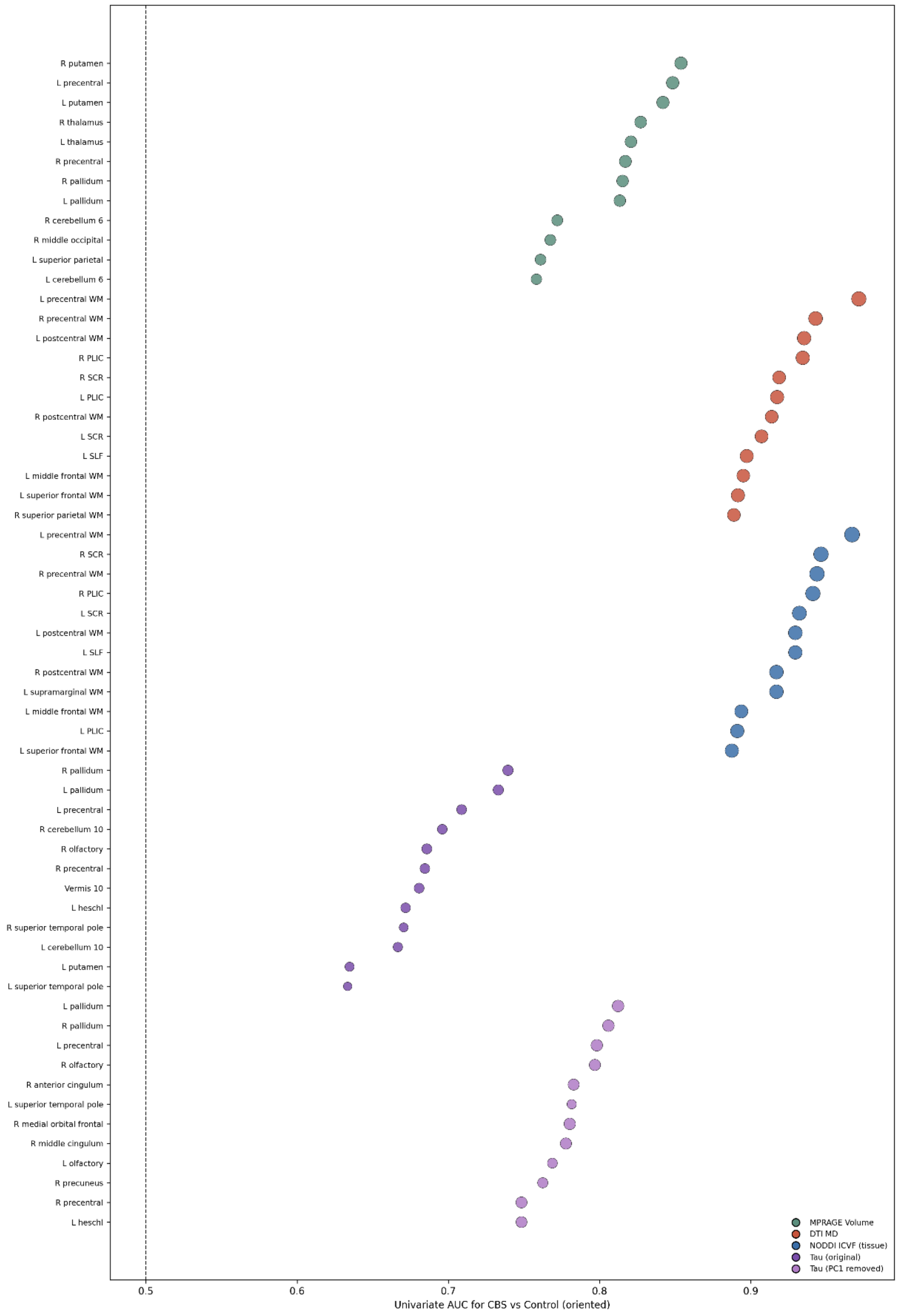

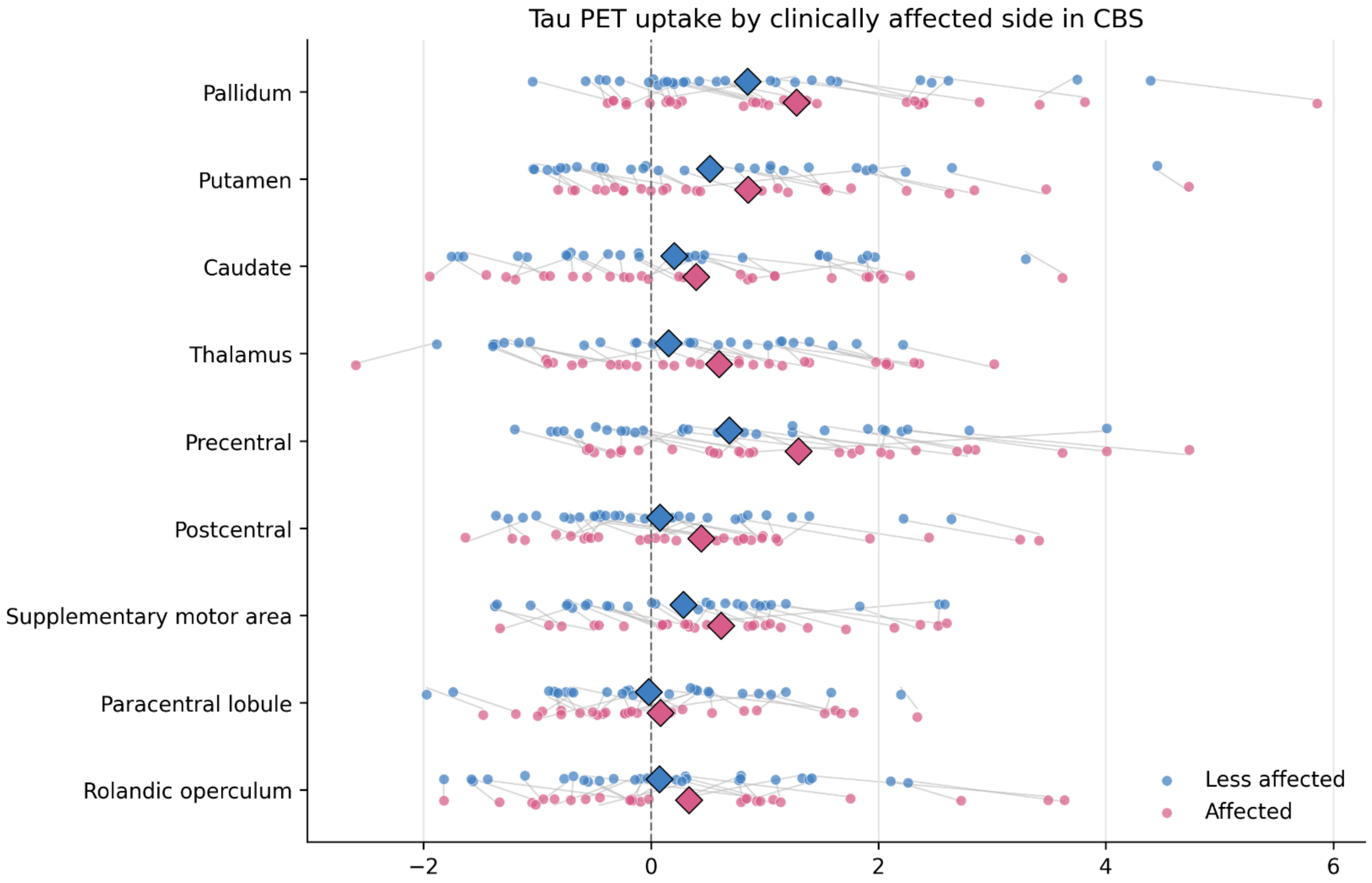


**Figure S2 Tau PET uptake in clinically affected and less affected hemisphere among CBS participants**. Hemispheres were aligned according to the clinically affected body side, with the affected brain hemisphere defined as the hemisphere contralateral to the more affected body side. Points show participant-level tau PET SUVR z-scores for selected motor and subcortical ROIs, lines connect paired hemispheric values within participants, and diamonds show group means. Z-scores were calculated relative to controls within each ROI.

**Table S4 The top 8 ROIs per modality distinguishing CBS from controls, sorted by AUC and effect size.**

| modality | rank | roi | AUC | Cohens_d | p_value | direction | sig |
|---|---|---|---|---|---|---|---|
| MPRAGE volume | 1 | R putamen | 0.854 | -1.47 | 1.83e-07 | CBS<Control | * |
| MPRAGE volume | 2 | L precentral | 0.848 | -1.49 | 2.34e-07 | CBS<Control | * |
| MPRAGE volume | 3 | L putamen | 0.842 | -1.45 | 3.42e-07 | CBS<Control | * |
| MPRAGE volume | 4 | R thalamus | 0.827 | -1.29 | 2.03e-06 | CBS<Control | * |
| MPRAGE volume | 5 | L thalamus | 0.821 | -1.25 | 6.16e-06 | CBS<Control | * |
| MPRAGE volume | 6 | R precentral | 0.817 | -1.35 | 1.84e-06 | CBS<Control | * |
| MPRAGE volume | 7 | R pallidum | 0.815 | -1.23 | 4.77e-06 | CBS<Control | * |
| MPRAGE volume | 8 | L pallidum | 0.813 | -1.22 | 5.6e-06 | CBS<Control | * |
| DTI MD | 1 | L precentral WM | 0.971 | 2.19 | 1.65e-10 | CBS>Control | * |
| DTI MD | 2 | R precentral WM | 0.943 | 1.97 | 2.54e-09 | CBS>Control | * |
| DTI MD | 3 | L postcentral WM | 0.935 | 1.86 | 1.03e-08 | CBS>Control | * |
| DTI MD | 4 | R PLIC | 0.934 | 1.87 | 2.26e-09 | CBS>Control | * |
| DTI MD | 5 | R SCR | 0.919 | 1.65 | 7.51e-08 | CBS>Control | * |
| DTI MD | 6 | L PLIC | 0.917 | 1.81 | 1.22e-08 | CBS>Control | * |
| DTI MD | 7 | R postcentral WM | 0.914 | 1.61 | 2.2e-07 | CBS>Control | * |
| DTI MD | 8 | L SCR | 0.907 | 1.67 | 5.58e-08 | CBS>Control | * |
| NODDI ICVF | 1 | L precentral WM | 0.967 | -2.39 | 3.96e-12 | CBS<Control | * |
| NODDI ICVF | 2 | R SCR | 0.946 | -2.25 | 1.93e-12 | CBS<Control | * |
| NODDI ICVF | 3 | R precentral WM | 0.944 | -2.30 | 7.14e-12 | CBS<Control | * |
| NODDI ICVF | 4 | R PLIC | 0.941 | -2.22 | 8.87e-12 | CBS<Control | * |
| NODDI ICVF | 5 | L SCR | 0.932 | -2.06 | 1.11e-10 | CBS<Control | * |
| NODDI ICVF | 6 | L postcentral WM | 0.929 | -1.94 | 5.92e-10 | CBS<Control | * |
| NODDI ICVF | 7 | L SLF | 0.929 | -1.85 | 4.42e-09 | CBS<Control | * |
| NODDI ICVF | 8 | R postcentral WM | 0.917 | -1.95 | 4.74e-10 | CBS<Control | * |
| Tau original | 1 | R pallidum | 0.739 | 0.85 | 0.00219 | CBS>Control | * |
| Tau original | 2 | L pallidum | 0.733 | 0.84 | 0.00248 | CBS>Control | * |
| Tau original | 3 | L precentral | 0.709 | 0.70 | 0.0105 | CBS>Control | * |
| Tau original | 4 | R cerebellum 10 | 0.696 | -0.67 | 0.0175 | CBS<Control | * |
| Tau original | 5 | R olfactory | 0.686 | -0.76 | 0.00619 | CBS<Control | * |
| Tau original | 6 | R precentral | 0.685 | 0.67 | 0.014 | CBS>Control | * |
| Tau original | 7 | Vermis 10 | 0.681 | -0.71 | 0.0115 | CBS<Control | * |
| Tau original | 8 | L heschl | 0.672 | -0.65 | 0.0179 | CBS<Control | * |
| Tau PC1-removed | 1 | L pallidum | 0.812 | 1.21 | 3.69e-05 | CBS>Control | * |

| | | | | | | | |
|---|---|---|---|---|---|---|---|
| Tau PC1-removed | 2 | R pallidum | 0.806 | 1.19 | 5.1e-05 | CBS>Control | * |
| Tau PC1-removed | 3 | L precentral | 0.798 | 1.19 | 5.42e-05 | CBS>Control | * |
| Tau PC1-removed | 4 | R olfactory | 0.797 | -1.09 | 0.000137 | CBS<Control | * |
| Tau PC1-removed | 5 | R anterior cingulum | 0.783 | -1.07 | 0.000193 | CBS<Control | * |
| Tau PC1-removed | 6 | L superior temporal pole | 0.782 | -0.56 | 0.0397 | CBS<Control | * |
| Tau PC1-removed | 7 | R medial orbital frontal | 0.780 | -1.17 | 5.03e-05 | CBS<Control | * |
| Tau PC1-removed | 8 | R middle cingulum | 0.778 | -1.09 | 0.000146 | CBS<Control | * |

**Table S5** Bootstrap comparison of sensorimotor summary AUCs. (A) AUCs were calculated for distinguishing CBS from controls using prespecified sensorimotor summary measures. (B) Pairwise AUC differences were estimated using stratified bootstrap resampling within diagnostic group with 10,000 iterations; p values are two-sided bootstrap p values and q values are Benjamini-Hochberg FDR-adjusted.

Post hoc amyloid-negative control restriction yielded similar sensorimotor AUC rankings: DTI MD 0.977, NODDI ICVF 0.920, GM volume 0.562, WM volume 0.730, PC1-removed tau 0.659, original tau 0.670.

**(A)**

| feature | label | control_mean | cbs_mean | auc | auc_ci_low | auc_ci_high |
|---|---|---|---|---|---|---|
| **motor_dti_md_z** | DTI MD | -1.82E-15 | 2.996 | 0.976 | 0.934 | 1.000 |
| **motor_icvf_tissue_z** | NODDI ICVF | 5.57E-16 | -2.266 | 0.935 | 0.864 | 0.988 |
| **gm_motor_volume_z** | GM volume | 6.03E-17 | -0.333 | 0.618 | 0.477 | 0.754 |
| **wm_motor_volume_z** | WM volume | -1.71E-16 | -0.802 | 0.767 | 0.642 | 0.880 |
| **motor_tau_pc1sub_z** | Tau PC1-removed | -4.40E-16 | 0.879 | 0.688 | 0.540 | 0.825 |
| **motor_tau_z** | Tau original | 3.29E-16 | 0.399 | 0.622 | 0.467 | 0.768 |

**(B)**

| feature_a | feature_b | auc a | auc b | auc_diff a-b | diff_ci_low | diff_ci_high | p_bootstrap_two_sided | q_bootstrap_bh |
|---|---|---|---|---|---|---|---|---|
| motor_dti_md_z | motor_icvf_tissue_z | 0.976 | 0.935 | 0.041 | 0.001 | 0.096 | 0.043 | 0.065 |
| motor_dti_md_z | gm_motor_volume_z | 0.976 | 0.618 | 0.358 | 0.220 | 0.500 | 0.000 | 0.000 |
| motor_dti_md_z | wm_motor_volume_z | 0.976 | 0.767 | 0.209 | 0.103 | 0.328 | 0.000 | 0.000 |
| motor_dti_md_z | motor_tau_pc1sub_z | 0.969 | 0.688 | 0.281 | 0.148 | 0.425 | 0.000 | 0.000 |
| motor_dti_md_z | motor_tau_z | 0.969 | 0.622 | 0.347 | 0.201 | 0.501 | 0.000 | 0.000 |

| | | | | | | | | |
|---|---|---|---|---|---|---|---|---|
| motor_icvf_tissue_z | gm_motor_volume_z | 0.935 | 0.618 | 0.317 | 0.171 | 0.462 | 0.000 | 0.000 |
| motor_icvf_tissue_z | wm_motor_volume_z | 0.935 | 0.767 | 0.168 | 0.051 | 0.294 | 0.003 | 0.005 |
| motor_icvf_tissue_z | motor_tau_pc1sub_z | 0.935 | 0.688 | 0.246 | 0.111 | 0.392 | 0.000 | 0.000 |
| motor_icvf_tissue_z | motor_tau_z | 0.935 | 0.622 | 0.313 | 0.152 | 0.475 | 0.000 | 0.001 |
| gm_motor_volume_z | wm_motor_volume_z | 0.618 | 0.767 | -0.149 | -0.246 | -0.056 | 0.001 | 0.002 |
| gm_motor_volume_z | motor_tau_pc1sub_z | 0.613 | 0.688 | -0.075 | -0.284 | 0.130 | 0.477 | 0.511 |
| gm_motor_volume_z | motor_tau_z | 0.613 | 0.622 | -0.009 | -0.217 | 0.204 | 0.932 | 0.932 |
| wm_motor_volume_z | motor_tau_pc1sub_z | 0.773 | 0.688 | 0.084 | -0.115 | 0.280 | 0.410 | 0.473 |
| wm_motor_volume_z | motor_tau_z | 0.773 | 0.622 | 0.151 | -0.047 | 0.342 | 0.136 | 0.185 |
| motor_tau_pc1sub_z | motor_tau_z | 0.688 | 0.622 | 0.066 | -0.087 | 0.213 | 0.392 | 0.473 |

### Amyloid-positive versus amyloid-negative CBS participants

**Table S6. ROI-level sensitivity analysis comparing amyloid-positive and amyloid-negative CBS participants.**The table shows the 15 ROI-level comparisons with the lowest Benjamini-Hochberg FDR-adjusted p values across dMRI and tau PET measures. No comparison survived FDR correction.

| modality | atlas | roi_label | abs_d | p_fdr_bh |
|---|---|---|---|---|
| DTI MD | JHU_TypeIII | ML L Medial lemniscus left | 1.210872794 | 0.082605241 |
| Tau (PC1 removed) | ADIR122 | Postcentral R | 1.036849049 | 0.086348855 |
| Tau (original) | ADIR122 | Frontal Inf Orb L | 0.942769466 | 0.088433451 |
| DTI MD | JHU_TypeIII | MFWM L MIDDLE FRONTAL WM left | 1.062294249 | 0.13407153 |
| NODDI ICVF (tissue) | JHU_TypeIII | EC L External capsule left | 1.1009217 | 0.13407153 |
| DTI MD | JHU_TypeIII | SCP L Superior cerebellar peduncle left | 1.118927337 | 0.13407153 |
| Tau (PC1 removed) | ADIR122 | Parietal Inf R | 1.258204419 | 0.13407153 |
| Tau (PC1 removed) | ADIR122 | Frontal Sup Orb R | 0.858450995 | 0.255843884 |
| DTI MD | JHU_TypeIII | EC L External capsule left | 1.22434766 | 0.255843884 |
| NODDI ICVF (tissue) | JHU_TypeIII | PTR L Posterior thalamic radiation left | 1.013546994 | 0.255843884 |
| Tau (PC1 removed) | ADIR122 | SupraMarginal R | 1.152549828 | 0.404709763 |
| Tau (original) | ADIR122 | Parietal Sup L | 0.701172513 | 0.419477825 |
| DTI MD | JHU_TypeIII | SCP R Superior cerebellar peduncle right | 1.245133506 | 0.419477825 |
| DTI MD | JHU_TypeIII | EC R External capsule right | 0.988946353 | 0.428451162 |

### Volume and Tau prediction of dMRI

**Table S7. Model comparison**

| n | outcome | predictors | r_squared | adj_r_squared | beta__motor_atrophy_score | p_value__motor_atrophy_score | model_name | beta__motor_tau_z_original | p_value__motor_tau_z_original | beta__motor_tau_z_pc1sub | p_value__motor_tau_z_pc1sub |
|---|---|---|---|---|---|---|---|---|---|---|---|
| **32** | motor_DTI_MD_mean_z | motor_atrophy_score | 0.387849 | 0.29716 | 0.995378 | 0.002579 | volume_only | | | | |
| **29** | motor_DTI_MD_mean_z | motor_tau_z_original | 0.138727 | -0.00482 | | | tau_original_only | 0.120112 | 0.707862 | | |
| **29** | motor_DTI_MD_mean_z | motor_atrophy_score + motor_tau_z_original | 0.389456 | 0.256729 | 1.046247 | 0.005377 | volume_plus_tau_original | -0.09196 | 0.746447 | | |
| **29** | motor_DTI_MD_mean_z | motor_tau_z_pc1sub | 0.233553 | 0.105812 | | | tau_pc1sub_only | | | 0.453903 | 0.089526 |
| **29** | motor_DTI_MD_mean_z | motor_atrophy_score + motor_tau_z_pc1sub | 0.389826 | 0.25718 | 0.946514 | 0.023463 | volume_plus_tau_pc1sub | | | 0.096189 | 0.730985 |
